# Religiosity and Spirituality as Stressors or Stress Relievers in relation to Sleep Health among African American Women

**DOI:** 10.1101/2025.08.22.25334167

**Authors:** Rupsha Singh, Symielle A. Gaston, Jason Ashe, Quaker E. Harmon, Yusuf Ransome, Ganesa Wegienka, Donna D. Baird, Harold G. Koenig, Chandra L. Jackson

## Abstract

**Objective:** To investigate associations between religiosity/spirituality and sleep and potential modification by stress among Black/African American (AA) women.

**Methods:** Using data from the Study of Environment, Lifestyle, & Fibroids at enrollment (2010– 2012) and three follow-up periods (2012–2018), we estimated prevalence ratios (PRs), risk ratios, and 95% confidence intervals (CIs) for the following sleep dimensions: short sleep duration (<7 hours), nonrestorative sleep (NRS, waking rested <4 days/week), and insomnia symptoms (difficulty falling/staying asleep). At baseline, participants reported: importance of faith, religion/spirituality as a source of strength/comfort (‘very-to-somewhat’ vs. ‘not at all’), and prayer/meditation frequency (‘everyday,’ ‘every week,’ or ‘≥once/month’ vs. ‘<once/month’). Day-to-day stress was dichotomized as ‘very high/moderate’ vs. ‘mild/not at all.’ PRs were estimated using adjusted Poisson regression with robust variance, and longitudinal models used generalized estimating equations. Interactions between religiosity/spirituality and stress were tested.

**Results:** Among 1,693 Black/AA women (mean age ± SD 29.2 ± 3.4 years), 69.3% reported faith as important, 55.6% perceived religion/spirituality as a source of strength/comfort, 58.8% prayed/meditated daily, and 43.8% reported high/moderate stress. Short sleep affected 58.4% of participants, NRS 9.5%, and insomnia symptoms 17.9%. Everyday vs. <once/month prayer/meditation was associated with NRS (aPR=3.16 [95% CI:1.16-8.56]). Among women experiencing daily life stress vs. no daily life stress, those who reported religion/spirituality as “very much” a source of strength/comfort had significantly lower prevalence of NRS compared to those who did not (aPR = 0.37, 95% CI: 0.14–0.98, p = 0.05). Religiosity/spirituality was not longitudinally associated with sleep.

**Conclusion:** Daily prayer/meditation was associated with NRS. Religion/spirituality as a source of strength was associated with restorative sleep among highly stressed women. Future research should address potential reverse causation for both negative and positive effects.

## INTRODUCTION

Poor sleep is of great public health concern across the globe.^1^ In the United States (US), variations in sleep health exist across racial, ethnic, and socioeconomic groups. For example, Black/African American (AA) adults are generally more likely than many other groups to experience short sleep duration (SSD), non-restorative sleep (NRS), and insomnia symptoms (IS).^2, 3^ These sleep disturbances can influence a broad array of health conditions over the lifespan by contributing to adverse health outcomes such as obesity, type 2 diabetes, cardiovascular disease, and mental health conditions such as depression.^4–6^

Multiple social and environmental stressors contribute to these preventable differences in sleep, including economic hardship, neighborhood, and workplace conditions.^7^ Within this broader context, Black/AA women face a distinct and compounded set of challenges due to the intersection of race, sex, and socioeconomic position.^8^ Stressors such as financial instability that contribute to housing and food insecurity, differential treatment in the workplace, caregiving responsibilities, socialization to be strong and invulnerable, and insufficient social support may heighten the social vulnerability of Black women.^9–11^ This cumulative and toxic stress, often referred to as “weathering,” exacerbates chronic conditions such as obesity, hypertension, and type 2 diabetes, which are bidirectionally associated with sleep disturbances.^12–15^

Religion and spirituality (R/S) may function as a coping mechanism in response to such stressors and is also a well-known independent determinant of health although evidence remains sparse for sleep health.^16–19^ According to the Religious Landscape Study conducted by the Pew Research Center, 80% of Black women have described religion as “very important” compared to 53% of the general U.S. population.^20^ Involving practices such as prayer, meditation, and church attendance, R/S has long been recognized as a culturally salient coping mechanism that fosters psychological resilience and provides social, emotional, informational, and financial support during adverse conditions.^19, 21^ Beyond its stress-buffering functions, R/S may directly shape health-related behaviors, regulate physiological functioning, and foster identity and community norms that influence both mental and physical health.^19^ For Black/AA women, R/S is often deeply intertwined with cultural traditions, gendered expectations, and the historical role of the Black church.^22–25^ Religious involvement may encompass active participation in women’s ministries, leadership in faith-based community outreach, and serving as a spiritual anchor for one’s family. The Black church has historically served not only as a religious institution but also as a center for social activism, civic engagement, and health promotion, creating pathways through which R/S influences both individual and community well-being.^22–26^ These intersecting cultural, social, and gendered roles may enhance the protective effects of R/S through strengthened social support networks, shared identity, and empowerment, but also may introduce stress when religious obligations compete with work, caregiving, and other life demands.^27^

Research indicates that perceptions of divine control and religious coping are associated with markers of resilience (i.e., higher adrenal hormone DHEAS and lower cortisol levels) among Black women.^28^ These physiological responses can directly influence sleep health by reducing hyperarousal and promoting relaxation. Such pathways may operate independently of external stress exposure by fostering routine, meaning-making, social cohesion, and health-promoting behaviors (e.g., avoiding substances, maintaining structure), in addition to reducing psychological distress and lowering allostatic load.^17, 19^ Moreover, religious involvement and coping have been associated with better mental and cardiovascular health among Black/AA adults.^29, 30^ A review of seven studies conducted by Hill et al. also found that practices such as prayer, meditation, and participation in faith-based communities have been associated with favorable sleep outcomes, including improved sleep quality, reduced sleep disturbances, and acquiring the recommended sleep duration.^17^

Nonetheless, the relationship between R/S and sleep may be complex and not universally positive.^18^ While R/S may act as a buffer against stress to promote restorative sleep, it may also introduce additional stress under certain conditions. For example, religious obligations or commitments may conflict with other responsibilities (e.g., work and family life), thereby creating psychological stress and disrupting sleep routines.^31^ Beyond time demands, normative expectations and fear of social sanction within faith communities (i.e., semi-involuntary participation) can create social-evaluative stress that also impairs sleep.^32^ This may be especially true for women in general and Black women, in particular, due to the historical and ongoing centrality of the church in African American communities, cultural expectations to serve as spiritual anchors for family and congregation, and the intersection of racialized and gendered stressors.^23, 33^ Individuals experiencing excessive life stress or major life challenges may respond by increasing their engagement in R/S activities, such as attending services more frequently, engaging in extended prayer, or participating in religious retreats, which can provide comfort and community support but may also place additional demands on time and energy that exacerbate stress and interfere with sleep.^34^ Similarly, individuals experiencing religious struggles/trauma, such as guilt, doubt, or judgement, may find that these experiences exacerbate stress and negatively affect sleep health.^35–38^ This bidirectionality highlights the need to consider the protective and potentially adverse effects of R/S on sleep health.

Important gaps persist in the literature. Much of the existing work is cross-sectional, limiting conclusions about directionality or causality, and often relies on narrow or unidimensional measures of religiosity (e.g., attendance only) and single-item or unvalidated sleep assessments.^17, 19, 28^ Although some studies provided biological plausibility by linking R/S to stress-related biomarkers,^28^ and others to mental and self-rated health,^29, 30^ few explicitly examined sleep health as a primary outcome or consider the interactive effects of R/S and psychosocial stress.^17, 19, 28^ Among the studies reviewed by Hill et al., none included Black women, all were conducted in the United States, and the majority involved Christian participants and middle-aged or older adults.^17^ Additionally, few studies considered potentially adverse aspects of religiosity, such as religious struggle.^17^ Collectively, these gaps underscore the need for rigorous, multidimensional, and population-specific research to better understand how R/S influences sleep health, particularly in the context of chronic stress among Black women.

This study builds on existing literature by examining stress as a potential modifier of the relationship between R/S and sleep health outcomes, including SSD, NRS, and IS, among Black/AA women. We hypothesized that 1) higher vs. lower R/S would be associated with lower prevalence and risk of SSD, NRS, and IS; and 2) daily life stress would modify the relationship between R/S and sleep, such that R/S may mitigate the adverse effects of stress on sleep in the presence of moderate/high stress but may have a less pronounced or potentially negative impact when day-to-day stress is absent or mild.

## METHODS

### Data Source: Study of Environmental, Lifestyle & Fibroids

The Study of Environmental, Lifestyle & Fibroids (SELF) is a prospective cohort study designed to identify risk factors for uterine fibroid incidence and growth among AA women. Enrolled participants were self-identified Black/AA (N = 1,693) women between the ages of 23-35 years who lived in Detroit, Michigan, and had no prior diagnosis of fibroids. The SELF protocol and a detailed list of exclusion and inclusion criteria have been published previously.^39^ Data were collected using self-administered computer-assisted web interviewing (CAWI) questionnaires and computer-assisted telephone interviews (CATI) at baseline (2010-2012), and during three follow-ups at approximately 20-month intervals. All participants provided written informed consent, and the SELF protocol was approved by the Institutional Review Board of the National Institute of the Environmental Health Sciences and Henry Ford Health System.

### Study Population

To be eligible for the current study, participants needed to have complete data on R/S (collected at baseline), and at least one sleep dimension (i.e., sleep duration, sleep quality, or insomnia symptoms). For the cross-sectional analyses, which examined baseline R/S in relation to baseline sleep, the analytic sample included all 1,693 Black/AA women with complete data on R/S and baseline sleep measures. For the longitudinal analyses, which examined baseline R/S in relation to repeated measures of sleep across baseline and three follow-up periods, the analytic sample also included all 1,693 Black/AA women; all available person-visit observations were used (6,062 for short sleep duration and 6,128 for non-restorative sleep and insomnia symptoms), with denominators differing due to visit-level missingness. Due to the small sample size (n=19), long sleep duration (>9 hours) was excluded from the analysis where sleep duration was the outcome.

### Exposure assessment: Religiosity and Spirituality

At baseline, participants self-reported importance of faith by responding to the question “How important is your religious faith or spirituality to you?” The responses were categorized as “not at all important,” “somewhat important”, “moderately important”, or “very important,” with “not at all important” serving as the reference category. Participants self-reported R/S as a source of strength and comfort by responding to the following question: “How much is religion or spirituality a source of strength and comfort to you?”. The responses were categorized as “not at all”, “somewhat”, “quite a bit”, or “very much,” with “not at all” serving as the reference category. Prayer frequency was defined by participant’s response to “How often do you pray or meditate?” The responses were categorized as “never/less than once a month” vs. “monthly or few days per month”, “weekly or a few days per week”, “everyday or several times per day,” with “never/less than once a month” serving as the reference category.

### Outcome assessment: Self-reported sleep duration, nonrestorative sleep (NRS), and insomnia symptoms (IS)

Participants provided self-reported data on sleep duration, NRS, and IS at baseline and during each follow-up assessment. Each measure was operationalized by categorizing responses in alignment with prior literature.^15, 40^ Sleep duration was assessed by asking, “How many hours of sleep do you usually get before a workday?” Based on the National Sleep Foundation categories, average sleep duration per day was defined as either short (<7 hours) or recommended (7–9 hours).^41^ NRS was measured by the frequency of waking up feeling rested, based on responses to the question, “How many days per week do you wake up feeling well rested?” Responses were dichotomized (<4 days/week vs. ≥4 days/week).^15, 40^ IS was assessed by asking, “About how many days per month do you have trouble falling asleep or going back to sleep after waking up?” Responses were dichotomized to indicate the presence of IS (≥10–14 days/month vs. 0–9 days/month).^15, 40^

### Potential Confounders

Potential confounding variables included sociodemographic and clinical characteristics that were selected a-priori based on the literature.^42, 43^ Sociodemographic variables were collected at baseline as well as each follow-up and included age, marital status, educational attainment, employment, and annual household income. Clinical characteristics included body mass index (BMI) (assessed with measured height and weight, kg/m^2^) and self-reported general health status at baseline and the three follow-ups.

### Potential Modifier

Daily life stress was assessed as a potential modifier by asking, “How stressful is your day-to-day life?” Responses included “very stressful”, “moderately stressful,” “mildly stressful,” and “not at all stressful”, which were then dichotomized as moderately/very stressful (high) vs. not at all/mildly stressful (low).

### Statistical Analysis

We computed descriptive statistics to summarize baseline sociodemographic characteristics, as well as health behaviors and characteristics, including sleep dimensions and daily stress. Age (treated as a continuous variable) which was summarized using mean and standard deviation; all other variables were categorical and summarized using frequencies and percentages. We compared baseline characteristics across categories of R/S dimensions, including importance of faith, R/S as a source of comfort/strength, and frequency of prayer/meditation. Statistical significance was assessed using one-way analysis of variance (ANOVA) for continuous variables and chi-squared tests for categorical variables, with p-values <0.05 considered statistically significant. We conducted cross-sectional analyses to estimate prevalence ratios (PRs) and 95% confidence intervals (CIs) for the associations between baseline R/S (i.e., importance of faith, R/S as a source of comfort and strength, and prayer/meditation frequency) and sleep dimensions, including sleep duration, NRS, and IS using Poisson regression with robust variance. We applied generalized estimating equations (GEE) to fit log-binomial regression models, estimating risk ratios (RRs) and 95% CIs for the longitudinal associations between baseline religiosity/spirituality and repeated measures of sleep duration, NRS, and IS (i.e., baseline to the third follow-up). All models were adjusted for baseline age, marital status, educational attainment, employment status, household income, BMI, and overall health status; visit-specific values were used in longitudinal analyses and baseline values in cross-sectional analyses. Effect modification by daily stress (high vs. low/no) was tested by including interaction terms between each R/S level and the visit-specific stress measure (i.e., stress assessed at the same visit as the sleep outcome) in the longitudinal models and baseline stress in the cross-sectional models. Statistical significance was defined as a two-sided p-value <0.05. All analyses were conducted using IBM SPSS version 29.

## RESULTS

### Study population characteristics

Among the 1,614 participants, the mean age was 29.2 with a standard deviation of 3.4 years. Baseline characteristics of the participants stratified by R/S are presented in Tables 1-3. R/S were commonly reported; 69.3% considered faith very important, 55.6% viewed religion or spirituality as a major source of strength or comfort, and 58.8% reported praying or meditating daily. Sleep disturbances were common, with 58.4% of women reporting SSD, 17.9% reporting IS, and 9.5% experiencing NRS. A high level of daily life stress was reported by 43.8% of participants.

**Table 1.** Baseline characteristics of participants by dimensions of importance of faith, Study of Environment, Lifestyle and Fibroids, N = 1693.

| Sociodemographic characteristics | Importance of Faith |  |  |  |  | p-value |
| --- | --- | --- | --- | --- | --- | --- |
|  | Not at all<br>N = 58 (3.4%) | Somewhat important<br>N = 181 (10.7%) | Moderately important<br>N = 280 (16.5%) | Very important<br>n = 1174 (69.3%) | Total<br>n = 1693 (100.0%) |  |
| Age, years (mean ± SD) | 28.4 ± 3.5 | 29.1 ± 3.4 | 28.3 ± 3.4 | 29.5 ± 3.4 | 29.2 ± 3.4 | <.001 |
| Educational attainment, n (%) |  |  |  |  |  |  |
| ≤High school | 23 (39.7) | 62 (34.3) | 73 (26.1) | 212 (18.1) | 370 (21.9) | <0.001 |
| Some college or Associate's/Technical degree | 22 (37.9) | 88 (48.6) | 138 (49.3) | 600 (51.1) | 848 (50.1) |  |
| ≥Bachelor's degree | 13 (22.4) | 31 (17.1) | 69 (24.6) | 362 (30.8) | 475 (28.1) |  |
| Employment status, n (%) |  |  |  |  |  |  |
| Not employed | 28 (48.3) | 88 (48.6) | 107 (38.2) | 417 (35.6) | 640 (37.9) | 0.025 |
| Employed part-time | 6 (10.3) | 18 (9.9) | 32 (11.4) | 154 (13.2) | 210 (12.4) |  |
| Employed full-time | 24 (41.4) | 75 (41.4) | 141 (50.4) | 600 (51.2) | 840 (49.7) |  |
| Annual household income, n (%) |  |  |  |  |  |  |
| <\$20,000 | 36 (62.1) | 102 (56.4) | 130 (46.4) | 505 (43.0) | 773 (45.7) | 0.002 |
| \$20,000 to \$50,000 | 15 (25.9) | 52 (28.7) | 112 (40.0) | 451 (38.4) | 630 (37.2) | |
| >\$50,000 | 7 (12.1) | 27 (14.9) | 38 (13.6) | 218 (18.6) | 290 (17.1) | |
| Marital status, n (%) |  |  |  |  |  |  |
| Never married | 48 (82.8) | 114 (63.0) | 169 (60.4) | 661 (56.3) | 992 (58.6) | <0.001 |
| Previously married or living as married | 6 (10.3) | 14 (7.7) | 35 (12.5) | 181 (15.4) | 236 (13.9) |  |
| Currently married or living as married | 4 (6.9) | 53 (29.3) | 76 (27.1) | 332 (28.3) | 465 (27.5) |  |
| Health behaviors and characteristics |  |  |  |  |  |  |
| Smoking status, n (%) |  |  |  |  |  |  |
| Never | 38 (65.5) | 114 (63.0) | 198 (70.7) | 895 (76.2) | 1245 (73.5) | <0.001 |
| Former | 1 (1.7) | 19 (10.5) | 16 (5.7) | 89 (7.6) | 125 (7.4) |  |
| Current | 19 (32.8) | 48 (26.5) | 66 (23.6) | 190 (16.2) | 323 (19.1) |  |
| Alcohol use, n (%) |  |  |  |  |  |  |
| None | 17 (29.3) | 37 (20.4) | 48 (17.1) | 399 (34.0) | 501 (29.6) | <0.001 |
| Moderate | 29 (50.0) | 94 (51.9) | 158 (56.4) | 580 (49.4) | 861 (50.9) |  |
| Heavy | 12 (20.7) | 50 (27.6) | 74 (26.4) | 195 (16.6) | 331 (19.6) |  |
| BMI (kg/m²), n (%) |  |  |  |  |  |  |
| Underweight/normal (<25) | 13 (22.4) | 32 (17.7) | 46 (16.4) | 244 (20.8) | 335 (19.8) | 0.463 |
| Overweight (25 - <30) | 8 (13.8) | 36 (19.9) | 58 (20.7) | 248 (21.1) | 350 (20.7) |  |
| Obese (≥30) | 37 (63.8) | 113 (62.4) | 176 (62.9) | 682 (58.1) | 1008 (59.5) |  |
| Sleep duration, n (%) |  |  |  |  |  |  |
| Short (<7 hours) | 33 (56.9) | 106 (58.6) | 168 (60.0) | 682 (58.1) | 989 (58.4) | 0.752 |
| Recommended (7-9 hours) | 25 (43.1) | 71 (39.2) | 110 (39.3) | 479 (40.8) | 685 (40.5) |  |
| Long (>9 hours) | 0 (0.0) | 4 (2.2) | 2 (0.7) | 13 (1.1) | 19 (1.1) |  |
| Insomnia symptoms, n (%) |  |  |  |  |  |  |
| Yes | 7 (12.1) | 34 (18.8) | 60 (21.4) | 202 (17.2) | 303 (17.9) | 0.241 |
| No | 51 (87.9) | 147 (81.2) | 220 (78.6) | 972 (82.8) | 1390 (82.1) |  |
| Non-restorative sleep, n (%) |  |  |  |  |  |  |
| Yes | 2 (3.4) | 15 (8.3) | 26 (9.3) | 117 (10.0) | 160 (9.5) | 0.376 |
| No | 56 (96.6) | 166 (91.7) | 254 (90.7) | 1057 (90.0) | 1533 (90.5) |  |
| General health, n (%) |  |  |  |  |  |  |
| Poor/Fair | 13 (22.4) | 32 (17.7) | 58 (20.7) | 178 (15.2) | 281 (16.6) | 0.083 |
| Good | 18 (31.0) | 78 (43.1) | 106 (37.9) | 440 (37.5) | 642 (37.9) |  |
| Very good/Excellent | 27 (46.6) | 71 (39.2) | 116 (41.4) | 556 (47.4) | 770 (45.5) |  |
| Daily life stress, n (%) |  |  |  |  |  |  |
| High | 25 (43.1) | 87 (48.1) | 127 (45.4) | 502 (42.8) | 741 (43.8) | 0.544 |
| Low/No | 33 (56.9) | 94 (51.9) | 153 (54.6) | 672 (57.2) | 952 (56.2) |  |

Note. Values are mean ± SD or n (%). p-values were obtained using one-way ANOVA for Age and Pearson's $\chi^2$ tests for all other (categorical) variables; tests were two-sided with $\alpha=0.05$ . Missing values were excluded pairwise.
Moderate alcohol use = 1-5 drinks on days when having alcohol; Heavy alcohol use = 6+ drinks on days when having alcohol
Missing data for employment: n = 3

**Table 2.** Baseline characteristics of participants by dimensions of amount religion/spirituality is a source of strength, Study of Environment, Lifestyle and Fibroids, N = 1693.

| Sociodemographic characteristics | Amount religion/spirituality is a source of strength/comfort |  |  |  |  | p-value |
| --- | --- | --- | --- | --- | --- | --- |
|  | Not at all<br>n = 63 (3.7%) | Somewhat<br>n = 293 (17.3%) | Quite a bit<br>n = 395 (23.3%) | Very much<br>n = 942 (55.6%) | Total<br>n = 1693 (100%) |  |
| Age, years (mean ± SD) | 28.22 ± 3.51 | 29.22 ± 3.54 | 28.96 ± 3.34 | 29.39 ± 3.43 | 29.22 ± 3.44 | 0.019 |
| Educational attainment, n (%) |  |  |  |  |  |  |
| ≤High school | 26 (41.3) | 91 (31.1) | 94 (23.8) | 159 (16.9) | 370 (21.9) | <0.001 |
| Some college or Associate's/Technical degree | 24 (38.1) | 141 (48.1) | 203 (51.4) | 480 (51.0) | 848 (50.1) |  |
| ≥Bachelor's degree | 13 (20.6) | 61 (20.8) | 98 (24.8) | 303 (32.2) | 475 (28.1) |  |
| Employment status, n (%) |  |  |  |  |  |  |
| Not employed | 33 (52.4) | 129 (44.0) | 148 (37.6) | 330 (35.1) | 640 (37.9) | 0.014 |
| Employed part-time | 6 (9.5) | 32 (10.9) | 41 (10.4) | 131 (13.9) | 210 (12.4) |  |
| Employed full-time | 24 (38.1) | 132 (45.1) | 205 (52.) | 479 (51.0) | 840 (49.7) |  |
| Annual household income, n (%) |  |  |  |  |  |  |
| <\$20,000 | 43 (68.3) | 155 (52.9) | 167 (42.3) | 408 (43.3) | 773 (45.7) | <0.001 |
| \$20,000 to \$50,000 | 13 (20.6) | 98 (33.4) | 165 (41.8) | 354 (37.6) | 630 (37.2) | |
| >\$50,000 | 7 (11.1) | 40 (13.7) | 63 (15.9) | 180 (19.1) | 290 (17.1) | |
| Marital status, n (%) |  |  |  |  |  |  |
| Never married | 51 (81.0) | 1743 (59.4) | 225 (57.0) | 542 (57.5) | 992 (58.6) | 0.002 |
| Previously married | 7 (11.1) | 30 (10.2) | 54 (13.7) | 145 (15.4) | 236 (13.9) |  |
| Currently married | 5 (7.9) | 89 (30.4) | 116 (29.4) | 255 (27.1) | 465 (27.5) |  |
| Health behaviors and characteristics |  |  |  |  |  |  |
| Smoking status, n (%) |  |  |  |  |  |  |
| Never | 43 (68.3) | 192 (65.5) | 272 (68.9) | 738 (78.3) | 1245 (73.5) | <0.001 |
| Former | 1 (1.6) | 26 (8.9) | 34 (8.6) | 64 (6.8) | 125 (7.4) |  |
| Current | 19 (30.2) | 75 (25.6) | 89 (22.5) | 140 (14.9) | 323 (19.1) |  |
| Alcohol use, n (%) |  |  |  |  |  |  |
| None | 20 (31.7) | 57 (19.5) | 86 (21.8) | 338 (35.9) | 501 (29.6) | <0.001 |
| Moderate | 32 (50.8) | 159 (54.3) | 215 (54.4) | 455 (48.3) | 861 (50.9) |  |
| Heavy | 11 (17.5) | 77 (26.3) | 94 (23.8) | 149 (15.8) | 331 (19.6) |  |
| Body Mass Index (kg/m²), n (%) |  |  |  |  |  |  |
| Underweight/normal (<25) | 16 (25.4) | 54 (18.4) | 69 (17.5) | 196 (20.8) | 335 (19.8) | 0.1.22 |
| Overweight (25 - <30) | 6 (9.5) | 62 (21.2) | 96 (24.3) | 186 (19.7) | 350 (20.7) |  |
| Obese (≥30) | 41 (65.1) | 177 (60.4) | 230 (58.2) | 560 (59.4) | 1008 (59.5) |  |
| Sleep duration, n (%) |  |  |  |  |  |  |
| Short (<7 hours) | 33 (52.4) | 180 (61.4) | 228 (57.7) | 548 (58.2) | 989 (58.4) | 0.788 |
| Recommended (7-9 hours) | 30 (47.6) | 110 (37.5) | 162 (41.0) | 383 (40.76) | 685 (40.5) |  |
| Long (>9 hours) | 0 (0.0) | 3 (1.0) | 5 (1.3) | 11 (1.2) | (1.1) |  |
| Insomnia symptoms, n (%) |  |  |  |  |  |  |
| Yes | 8 (12.7) | 67 (22.9) | 69 (17.5) | 159 (16.9) | 303 (17.9) | 0.079 |
| No | 55 (87.3) | 226 (77.1) | 326 (82.5) | 783 (83.1) | 1390 (82.1) |  |
| Non-restorative sleep, n (%) |  |  |  |  |  |  |
| Yes | 7 (11.1) | 19 (6.5) | 26 (6.6) | 108 (11.5) | 160 (9.5) | 0.009 |
| No | 56 (88.9) | 274 (93.5) | 369 (93.4) | 834 (88.5) | 1533 (90.5) |  |
| General health, n (%) |  |  |  |  |  |  |
| Poor/Fair | 12 (19) | 56 (19.1) | 83 (21.0) | 130 (13.8) | 281 (16.6) | <0.001 |
| Good | 21 (33.3) | 128 (43.7) | 148 (37.5) | 345 (36.6) | 642 (37.9) |  |
| Very good/Excellent | 30 (47.6) | 109 (37.2) | 164 (41.5) | 467 (49.6) | 770 (45.5) |  |
| Daily life stress, n (%) |  |  |  |  |  |  |
| High | 24 (38.1) | 135 (46.1) | 190 (48.1) | 392 (41.6) | 741 (43.8) | 0.100 |
| Low/No | 39 (61.9) | 158 (53.9) | 205 (51.9) | 550 (58.4) | 952 (56.2) |  |

Note. Values are mean ± SD or n (%). p-values were obtained using one-way ANOVA for Age and Pearson's $\chi^2$ tests for all other (categorical) variables; tests were two-sided with $\alpha=0.05$ . Missing values were excluded pairwise.
Moderate alcohol use = 1-5 drinks on days when having alcohol; Heavy alcohol use = 6+ drinks on days on having alcohol
Missing data for employment: n = 3

**Table 3.** Baseline characteristics of participants by dimensions of prayer/meditation frequency, Study of Environment, Lifestyle and Fibroids, N = 1693.

| Sociodemographic characteristics | Frequency of prayer/meditation |  |  |  |  | p-value |
| --- | --- | --- | --- | --- | --- | --- |
|  | Never/<once a month<br>n = 156 (9.2%) | ≥Once per month<br>n = 187 (11.0%) | Every week<br>n = 355 (21.0%) | Everyday<br>n = 995 (58.8%) | Total<br>n = 1693 (100%) |  |
| Age, years (mean ± SD) | 28.79 ± 3.58 | 28.77 ± 3.42 | 29.08 ± 3.29 | 29.42 ± 3.47 | 29.22 ± 3.44 | 0.021 |
| Educational attainment, n (%) |  |  |  |  |  |  |
| ≤High school | 64 (41.0) | 44 (23.5) | 57 (16.1) | 205 (20.6) | 370 (21.9) | <0.001 |
| Some college or Associate's/Technical degree | 62 (39.7) | 97 (51.9) | 186 (52.4) | 503 (50.6) | 848 (50.1) |  |
| ≥Bachelor's degree | 30 (19.2) | 46 (24.6) | 112 (31.5) | 287 (28.8) | 475 (28.1) |  |
| Employment status, n (%) |  |  |  |  |  |  |
| Not employed | 78 (50.0) | 76 (40.9) | 121 (34.1) | 365 (36.8) | 640 (37.9) | 0.012 |
| Employed part-time | 15 (9.6) | 15 (8.1) | 47 (13.2) | 133 (13.4) | 210 (12.4) |  |
| Employed full-time | 63 (40.4) | 95 (51.1) | 187 (52.7) | 495 (49.8) | 840 (49.7) |  |
| Annual household income, n (%) |  |  |  |  |  |  |
| <\$20,000 | 91 (58.3) | 88 (47.1) | 144 (40.6) | 450 (45.2) | 773 (45.7) | 0.002 |
| \$20,000 to \$50,000 | 39 (25.0) | 78 (41.7) | 147 (41.4) | 366 (36.8) | 630 (37.2) | |
| >\$50,000 | 26 (16.7) | 21 (11.2) | 64 (18.0) | 179 (18.0) | 290 (17.1) | |
| Marital status, n (%) |  |  |  |  |  |  |
| Never married | 116 (74.4) | 118 (63.1) | 208 (58.6) | 550 (55.3) | 992 (58.6) | <0.001 |
| Previously married | 14 (9.0) | 22 (11.8) | 45 (12.7) | 155 (15.6) | 236 (13.9) |  |
| Currently married | 26 (16.7) | 47 (25.1) | 102 (28.71) | 290 (29.1) | 465 (27.5) |  |
| Health behaviors and characteristics |  |  |  |  |  |  |
| Smoking status, n (%) |  |  |  |  |  |  |
| Never | 104 (66.7) | 138 (73.8) | 262 (73.8) | 741 (74.5) | 1245 (73.5) | 0.424 |
| Former | 11 (7.1) | 15 (8.0) | 26 (7.3) | 73 (7.3) | 125 (7.4) |  |
| Current | 41 (26.3) | 34 (18.2) | 67 (18.9) | 181 (18.2) | 323 (19.1) |  |
| Alcohol use, n (%) |  |  |  |  |  |  |
| None | 41 (26.3) | 42 (22.5) | 76 (21.4) | 342 (34.4) | 501 (29.6) | <0.001 |
| Moderate | 73 (46.8) | 99 (52.9) | 202 (56.9) | 487 (48.9) | 861 (50.9) |  |
| Heavy | 42 (26.9) | 46 (24.6) | 77 (21.7) | 166 (16.7) | 331 (19.6) |  |
| BMI (kg/m²), n (%) |  |  |  |  |  |  |
| Underweight/Normal(<25) | 35 (22.4) | 38 (20.3) | 60 (16.9) | 202 (20.3) | 335 (19.8) | 0.158 |
| Overweight (25 - <30) | 21 (13.5) | 45 (24.1) | 72 (20.3) | 212 (21.3) | 350 (20.7) |  |
| Obese (≥30) | 100 (64.1) | 104 (55.6) | 223 (62.8) | 581 (58.4) | 1008 (59.5) |  |
| Sleep duration, n (%) |  |  |  |  |  |  |
| Short (<7 hours) | 87 (55.8%) | 105 (56.1%) | 214 (60.3%) | 583 (58.6%) | 989 (58.4%) |  |
| Recommended (7-9 hours) | 68 (43.6%) | 78 (41.7%) | 138 (38.9%) | 401 (40.3%) | 685 (40.5%) | 0.739 |
| Long (>9 hours) | 1 (0.6%) | 4 (2.1%) | 3 (0.8%) | 11 (1.1%) | 19 (1.1%) |  |
| Insomnia symptoms, n (%) |  |  |  |  |  |  |
| Yes | 19 (12.2%) | 35 (18.7%) | 58 (16.3%) | 191 (19.2%) | 303 (17.9%) | 0.152 |
| No | 137 (87.8%) | 152 (81.3%) | 297 (83.7%) | 804 (80.8%) | 1390 (82.1%) |  |
| Non-restorative sleep, n (%) |  |  |  |  |  |  |
| Yes | 10 (6.4) | 19 (10.2) | 29 (8.2) | 102 (10.3) | 160 (9.5) | 0.359 |
| No | 146 (93.6) | 168 (89.8) | 326 (91.8) | 893 (89.7) | 1533 (90.5) |  |
| General health, n (%) |  |  |  |  |  |  |
| Poor/Fair | 34 (21.8) | 29 (15.5) | 57 (16.1) | 161 (16.2) | 281 (16.6) | 0.240 |
| Good | 62 (39.7) | 77 (41.2) | 143 (40.3) | 360 (36.2) | 642 (37.9) |  |
| Very good/Excellent | 60 (38.5) | 80 (43.3) | 155 (43.7) | 474 (47.6) | 770 (45.5) |  |
| Daily life stress, n (%) |  |  |  |  |  |  |
| High | 68 (43.6) | 78 (41.7) | 166 (46.8) | 429 (43.1) | 741 (43.8) | 0.618 |
| Low/No | 88 (56.4) | 109 (58.03) | 189 (53.2) | 566 (56.9) | 952 (56.2) |  |

Note. Values are mean ± SD or n (%). p-values were obtained using one-way ANOVA for Age and Pearson's $\chi^2$ tests for all other (categorical) variables; tests were two-sided with $\alpha=0.05$ . Missing values were excluded pairwise.
Moderate alcohol use = 1-5 drinks on days when having alcohol; Heavy alcohol use = 6+ drinks on days on having alcohol
Missing data for employment: n = 3

Compared with women who reported lower levels of R/S, those reporting higher R/S were generally older, had higher educational attainment and income, were more likely to be employed full-time, and were more often currently or previously married. Additionally, participants with higher R/S more frequently reported no smoking or alcohol use. Across all three R/S dimensions, women who reported higher R/S tended to report shorter sleep durations, although these differences were not statistically significant. IS did not significantly differ by the importance of faith (*p* = 0.24), source of strength/comfort (*p* = 0.08), or frequency of prayer/meditation (*p* = 0.15), although the proportion of participants reporting IS was slightly lower among those who considered faith very important (17.2% vs. 21.4% among those who considered faith moderately important), among those who considered R/S is very much a source of strength/comfort (16.9% vs. 22.9% among those who considered R/S somewhat a source of strength/comfort), and among those who prayed or meditated daily (19.2% vs. 16.3% among those who prayed every week). NRS differed in terms of statistical significance by strength/comfort from R/S (*p* = 0.01), with lowest prevalence among those who reported somewhat strength/comfort (6.5%). In contrast, no statistically significant differences were observed in NRS by importance of faith (*p* = 0.38) or prayer/meditation frequency (*p* = 0.36), although NRS tended to be less common among those who reported higher R/S engagement.

Daily life stress appeared more prevalent among those who considered faith somewhat important (48.1%) compared with those who considered it very important (42.8% among those reporting very important). Daily stress was more common among those who viewed religion or spirituality as very much a source of strength/comfort (41.6%) compared to those reporting not at all a strength/comfort (38.1%), as well as among those who prayed or meditated every week (46.8%) vs. those who prayed/meditated ≥once a month (41.7%). However, these differences were not statistically significant.

### Cross-sectional associations between R/S and sleep

Table 4 presents the adjusted cross-sectional associations between baseline R/S and sleep dimensions. None of the R/S indicators were significantly associated with SSD. Everyday prayer/meditation was associated with higher prevalence of NRS (adjusted prevalence ratios [aPR] = 3.16, 95% CI: 1.16–8.56, *p* = .02) compared to those who never engaged in prayer/meditation. No statistically significant associations were observed between R/S and IS.

**Table 4.** Adjusted prevalence ratios (aPRs) and 95% confidence intervals (CIs) for association between religiosity/spirituality and sleep at baseline, N = 1693.

| **Short sleep duration** |  |  |  |  |
| --- | --- | --- | --- | --- |
|  | **aPRs** | **95% CI** |  | **p-value** |
|  |  | **Lower** | **Upper** |  |
| Importance of faith |  |  |  |  |
| Very important | 1.00 | 0.72 | 1.38 | 0.97 |
| Moderately important | 1.11 | 0.78 | 1.56 | 0.57 |
| Somewhat important | 0.94 | 0.64 | 1.37 | 0.94 |
| Religion/Spirituality is a source of strength/support |  |  |  |  |
| Very much | 0.98 | 0.74 | 1.31 | 0.98 |
| Quite a bit | 0.90 | 0.66 | 1.23 | 0.51 |
| Somewhat | 1.05 | 0.77 | 1.42 | 0.78 |
| Frequency of prayer/meditation |  |  |  |  |
| Everyday | 1.07 | 0.86 | 1.34 | 0.55 |
| Every week | 1.16 | 0.91 | 1.48 | 0.22 |
| ≥Once per month | 0.92 | 0.68 | 1.23 | 0.56 |
| **Non-restorative sleep** |  |  |  |  |
|  | **aPRs** | **95% CI** |  | **p-value** |
|  |  | **Lower** | **Upper** |  |
| Importance of faith |  |  |  |  |
| Very important | 4.92 | 0.74 | 32.63 | 0.10 |
| Moderately important | 4.73 | 0.69 | 32.57 | 0.12 |
| Somewhat important | 3.84 | 0.53 | 27.72 | 0.18 |
| Religion/Spirituality as a source of strength/support |  |  |  |  |
| Very much | 2.92 | 0.77 | 11.08 | 0.12 |
| Quite a bit | 2.19 | 0.56 | 8.69 | 0.26 |
| Somewhat | 1.77 | 0.43 | 7.28 | 0.43 |
| Frequency of prayer/meditation |  |  |  |  |
| Everyday | **3.16** | **1.16** | **8.56** | **0.02** |
| Every week | 2.20 | 0.76 | 6.39 | 0.15 |
| ≥Once per month | 2.52 | 0.84 | 7.60 | 0.10 |
| **Insomnia symptoms** |  |  |  |  |
|  | **aPRs** | **95% CI** |  | **p-value** |
|  |  | **Lower** | **Upper** |  |
| Importance of faith |  |  |  |  |
| Very important | 0.84 | 0.33 | 2.12 | 0.70 |
| Moderately important | 1.15 | 0.43 | 3.07 | 0.78 |
| Somewhat important | 1.20 | 0.44 | 3.31 | 0.72 |
| Religion/Spirituality as a source of strength/support |  |  |  |  |
| Very much | 0.80 | 0.34 | 1.87 | 0.61 |
| Quite a bit | 0.87 | 0.35 | 2.12 | 0.75 |
| Somewhat | 1.20 | 0.50 | 2.92 | 0.68 |
| Frequency of prayer/meditation |  |  |  |  |
| Everyday | 1.21 | 0.63 | 2.33 | 0.57 |
| Every week | 1.14 | 0.55 | 2.35 | 0.73 |
| ≥Once per month | 1.10 | 0.49 | 2.48 | 0.82 |

Referent categories: importance of faith = “not at all important”; religion/spirituality as a source of strength/support = “not at all”; frequency of prayer/meditation = “never/less than once a month”
Models adjusted for baseline age, marital status, educational attainment, employment status, household income, body mass index (with Class I, II, and III obesity combined), and general health status.

### Longitudinal associations between R/S and sleep

Table 5 presents the adjusted longitudinal associations between baseline R/S and repeated measures of sleep across baseline and three follow-up assessments. None of the R/S indicators were statistically significantly associated with SSD, NRS, or IS over time.

**Table 5.** Adjusted risk ratios (aRRs) and 95% CIs for association between baseline religiosity/spirituality and repeated sleep (i.e., baseline to follow-up 3), N = 1693.

| **Short sleep duration** |  |  |  |  |
| --- | --- | --- | --- | --- |
|  | **aRRs** | **95% CI** |  | **p-value** |
|  |  | **Lower** | **Upper** |  |
| Importance of faith |  |  |  |  |
| Very important | 1.03 | 0.85 | 1.24 | 0.78 |
| Moderately important | 1.06 | 0.87 | 1.29 | 0.57 |
| Somewhat important | 1.01 | 0.81 | 1.25 | 0.95 |
| Religion/Spirituality as a source of strength/support |  |  |  |  |
| Very much | 1.09 | 0.89 | 1.33 | 0.40 |
| Quite a bit | 1.11 | 0.90 | 1.36 | 0.34 |
| Somewhat | 1.10 | 0.89 | 1.36 | 0.39 |
| Frequency of prayer/meditation |  |  |  |  |
| Everyday | 1.00 | 0.89 | 1.13 | 0.99 |
| Every week | 1.05 | 0.92 | 1.20 | 0.45 |
| ≥Once per month | 0.95 | 0.82 | 1.11 | 0.54 |
| **Non-restorative sleep** |  |  |  |  |
|  | **aRRs** | **95% CI** |  | **p-value** |
|  |  | **Lower** | **Upper** |  |
| Importance of faith |  |  |  |  |
| Very important | 0.93 | 0.77 | 1.13 | 0.48 |
| Moderately important | 1.05 | 0.85 | 1.29 | 0.65 |
| Somewhat important | 1.08 | 0.87 | 1.34 | 0.51 |
| Religion/Spirituality as a source of strength/support |  |  |  |  |
| Very much | 0.93 | 0.78 | 1.11 | 0.41 |
| Quite a bit | 1.00 | 0.83 | 1.21 | 0.97 |
| Somewhat | 1.00 | 0.83 | 1.21 | 0.98 |
| Frequency of prayer/meditation |  |  |  |  |
| Everyday | 0.91 | 0.81 | 1.03 | 0.14 |
| Every week | 0.89 | 0.77 | 1.03 | 0.11 |
| ≥Once per month | 0.99 | 0.85 | 1.15 | 0.88 |
| **Insomnia symptoms** |  |  |  |  |
|  | **aRRs** | **95% CI** |  | **p-value** |
|  |  | **Lower** | **Upper** |  |
| Importance of faith |  |  |  |  |
| Very important | 0.67 | 0.42 | 1.07 | 0.09 |
| Moderately important | 0.70 | 0.41 | 1.17 | 0.17 |
| Somewhat important | 0.71 | 0.41 | 1.23 | 0.23 |
| Religion/Spirituality as a source of strength/support |  |  |  |  |
| Very much | 0.87 | 0.50 | 1.50 | 0.61 |
| Quite a bit | 0.78 | 0.44 | 1.38 | 0.39 |
| Somewhat | 0.91 | 0.51 | 1.63 | 0.76 |
| Frequency of prayer/meditation |  |  |  |  |
| Everyday | 1.04 | 0.71 | 1.52 | 0.86 |
| Every week | 1.07 | 0.71 | 1.63 | 0.75 |
| ≥Once per month | 1.10 | 0.69 | 1.75 | 0.69 |

Note. Due to missingness, 6062 observations among 1693 participants were included in models for short sleep duration, and 6128 observations among 1693 participants were included in models for the remaining of sleep parameters.
Referent categories: importance of faith = “not at all important”; religion/spirituality as a source of strength/support = “not at all”; frequency of prayer/meditation = “never/less than once a month” Models adjusted for time-varying age, marital status, educational attainment, employment status, household income, body mass index (with Class I, II, and III obesity combined), and general health status.

### Modification of the R/S-Sleep relationship by Stress

Table 6 presents the results from cross-sectional models examining whether daily life stress modifies the associations between R/S and sleep dimensions. Interactions between daily life stress and R/S indicators were not statistically significantly associated with SSD. Similarly, there was no significant interaction between prayer/meditation frequency and daily life stress for any of the sleep outcomes. However, interactions that approached statistical significance emerged between daily life stress and the extent to which participants viewed R/S as a source of strength/comfort (p-value for interaction term = 0.059). Among women experiencing daily life stress vs. low/no daily life stress, those who reported R/S as “very much” (aPR = 0.37, 95% CI: 0.14–0.98, *p* = 0.05), “quite a bit” (aPR = 0.17, 95% CI: 0.05–0.59, *p* = 0.01), or “somewhat” (aPR = 0.26, 95% CI: 0.08–0.92, *p* = 0.04) a source of strength had significantly lower prevalence of NRS compared with those who did not derive strength/comfort from R/S. No statistically significant interactions were found between R/S indicators and daily life stress in relation to IS.

**Table 6.** Adjusted prevalence ratios (aPRs) and 95% confidence intervals (CIs) for association between religiosity/spirituality and sleep at baseline stratified by daily stress, N = 1693.

|  | aPRs | 95% CI |  | p-value | aPRs | 95% CI |  | p-value | Interaction<br>p-value |
| --- | --- | --- | --- | --- | --- | --- | --- | --- | --- |
| Daily Stress | High |  |  |  | Low/No |  |  |  |  |
| Short sleep duration |  |  |  |  |  |  |  |  |  |
| Importance of faith |  |  |  |  |  |  |  |  |  |
| Very important | 1.09 | 0.81 | 1.45 | 0.59 | 1.01 | 0.73 | 1.40 | 0.95 | 0.25 |
| Moderately important | 1.01 | 0.74 | 1.38 | 0.95 | 1.12 | 0.80 | 1.58 | 0.51 |  |
| Somewhat important | 1.13 | 0.82 | 1.55 | 0.45 | 0.94 | 0.65 | 1.37 | 0.76 |  |
| Religion/Spirituality is a source of strength/support |  |  |  |  |  |  |  |  |  |
| Very much | 1.35 | 0.91 | 2.01 | 0.14 | 1.00 | 0.75 | 1.33 | 1.00 | 0.29 |
| Quite a bit | 1.36 | 0.91 | 2.04 | 0.13 | 0.92 | 0.67 | 1.25 | 0.57 |  |
| Somewhat | 1.34 | 0.89 | 2.02 | 0.16 | 1.06 | 0.78 | 1.43 | 0.73 |  |
| Frequency of prayer/meditation |  |  |  |  |  |  |  |  |  |
| Everyday | 1.06 | 0.88 | 1.28 | 0.54 | 1.08 | 0.87 | 1.36 | 0.48 | 0.12 |
| Every week | 1.02 | 0.83 | 1.26 | 0.83 | 1.18 | 0.93 | 1.51 | 0.18 |  |
| ≥Once per month | 1.16 | 0.93 | 1.44 | 0.19 | 0.92 | 0.69 | 1.24 | 0.60 |  |
| Non-restorative sleep |  |  |  |  |  |  |  |  |  |
| Importance of faith |  |  |  |  |  |  |  |  |  |
| Very important | 1.19 | 0.16 | 8.66 | 0.87 | 5.20 | 0.80 | 33.91 | 0.09 | 0.80 |
| Moderately important | 1.46 | 0.18 | 11.74 | 0.72 | 4.62 | 0.68 | 31.31 | 0.12 |  |
| Somewhat important | 1.52 | 0.18 | 12.75 | 0.70 | 3.79 | 0.53 | 26.98 | 0.18 |  |
| Religion/Spirituality as a source of strength/support |  |  |  |  |  |  |  |  |  |
| Very much | **0.37** | **0.14** | **0.98** | **0.05** | 3.17 | 0.84 | 11.97 | 0.09 | 0.06 |
| Quite a bit | **0.17** | **0.05** | **0.59** | **0.01** | 2.24 | 0.57 | 8.78 | 0.25 |  |
| Somewhat | **0.26** | **0.08** | **0.92** | **0.04** | 1.80 | 0.44 | 7.39 | 0.41 |  |
| Frequency of prayer/meditation |  |  |  |  |  |  |  |  |  |
| Everyday | 0.55 | 0.22 | 1.35 | 0.19 | 3.37 | 1.23 | 9.22 | 0.02 | 0.07 |
| Every week | 0.62 | 0.23 | 1.68 | 0.35 | 2.37 | 0.81 | 6.94 | 0.12 |  |
| ≥Once per month | 0.94 | 0.33 | 2.69 | 0.91 | 2.63 | 0.87 | 7.96 | 0.09 |  |
| Insomnia symptoms |  |  |  |  |  |  |  |  |  |
| Importance of faith |  |  |  |  |  |  |  |  |  |
| Very important | 2.19 | 0.77 | 6.26 | 0.14 | 0.81 | 0.31 | 2.14 | 0.67 | 0.29 |
| Moderately important | 2.48 | 0.84 | 7.31 | 0.10 | 1.15 | 0.41 | 3.19 | 0.79 |  |
| Somewhat important | 1.84 | 0.61 | 5.55 | 0.28 | 1.20 | 0.42 | 3.42 | 0.73 |  |
| Religion/Spirituality as a source of strength/support |  |  |  |  |  |  |  |  |  |
| Very much | 2.01 | 0.70 | 5.79 | 0.20 | 0.77 | 0.32 | 1.86 | 0.56 |  |
| Quite a bit | 1.18 | 0.62 | 5.30 | 0.28 | 0.87 | 0.34 | 2.19 | 0.76 | 0.50 |
| Somewhat | 2.29 | 0.79 | 6.69 | 0.13 | 1.20 | 0.48 | 2.99 | 0.70 |  |
| Frequency of prayer/meditation |  |  |  |  |  |  |  |  |  |
| Everyday | 1.91 | 1.05 | 3.50 | 0.04 | 1.19 | 0.61 | 2.32 | 0.61 | 0.64 |
| Every week | 1.47 | 0.77 | 2.84 | 0.25 | 1.11 | 0.54 | 2.30 | 0.78 |  |
| ≥Once per month | 1.98 | 1.01 | 3.90 | 0.05 | 1.09 | 0.48 | 2.45 | 0.84 |  |

Referent categories: importance of faith = “not at all important”; religion/spirituality as a source of strength/support = “not at all”; frequency of prayer/meditation = “never/less than once a month”
Models adjusted for baseline age, marital status, educational attainment, employment status, household income, body mass index (with Class I, II and III obesity combined), and general health status.
Bolded estimates represent within-stratum associations statistically significant at $p < 0.05$ .

Table 7 presents the findings for statistical interactions between daily life stress and R/S for longitudinal associations. No interactions between R/S and daily life stress were statistically significant in any of the longitudinal models.

**Table 7.**
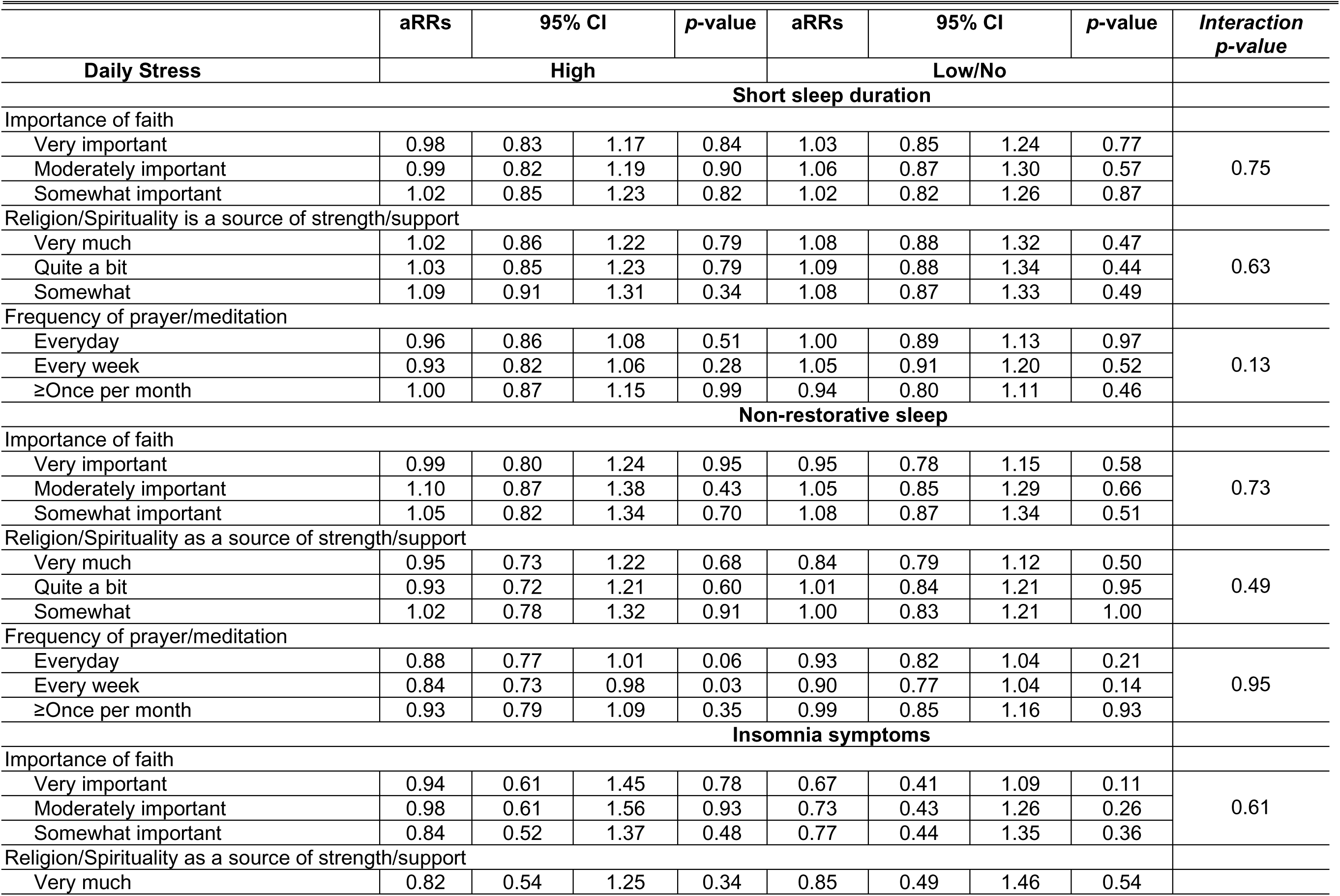
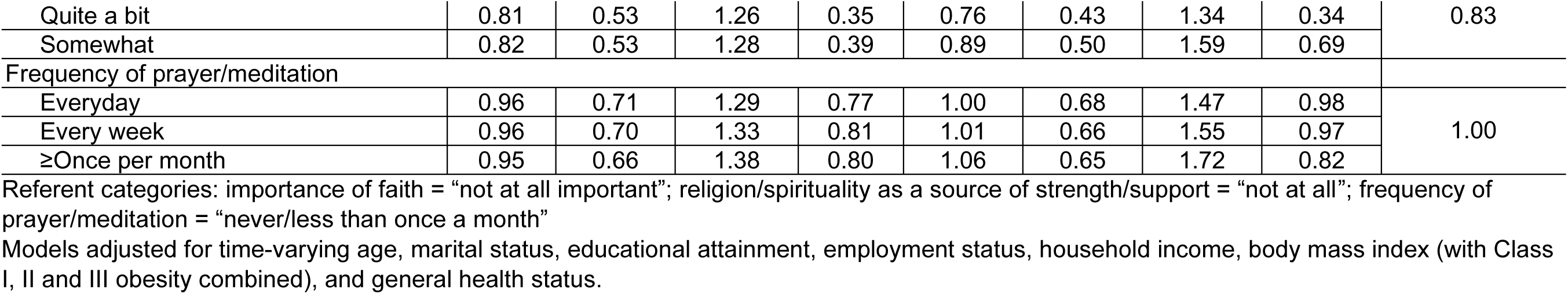
Adjusted risk ratios (aRRs) and 95% CIs for association between baseline religiosity/spirituality and repeated sleep (i.e., baseline to follow-up 3), stratified by daily stress, N = 1693.

## DISCUSSION

This study investigated cross-sectional and longitudinal associations between multiple dimensions of R/S and sleep health among young Black/AA women, and potential modification of this association by daily stress. Women in the SELF study reported a high importance of faith, frequently engaged in prayer or meditation, and often perceived R/S as a source of strength and comfort. SSD, IS, NRS, and daily life stress were also common among women in the study. We found that R/S was differentially associated with sleep health depending on the type of R/S indicator and the level of stress. Specifically, viewing R/S as a source of strength or comfort was marginally associated with a higher prevalence of restorative sleep among women reporting very high or moderate levels of day-to-day stress. In contrast, everyday prayer or meditation was associated with a higher prevalence of NRS in cross-sectional analyses. No significant associations were observed between R/S and sleep health over time in the longitudinal analysis.

These findings partially align with prior research suggesting that R/S can serve as a protective factor for sleep health, particularly in the context of chronic stress. For example, studies have shown that religious coping is associated with lower psychological distress, improved cardiovascular profiles, and reduced allostatic load, which may support sleep regulation through stress-buffering mechanisms.^17, 28–30^ Faith-based practices and beliefs may also offer meaning, belonging, and tools for coping with adversity. Among Black/AA women, R/S is often deeply integrated into cultural and community life, and has been shown to promote psychological resilience and emotional regulation.^19, 20, 29^ Our finding that R/S as a source of strength was associated with better sleep under stress is consistent with the stress-buffering hypothesis, which posits that the health benefits of psychosocial resources such as R/S are most pronounced when individuals are exposed to stressors, as the coping resource helps to mitigate stress-related physiological or emotional wear and tear.^44, 45^

Conversely, the finding that frequent prayer or meditation was associated with NRS may reflect situations where individuals engage in religious coping in response to existing distress, including poor sleep. This possibility highlights the complex and potentially bidirectional nature of the relationship between R/S and sleep, including the potential for reverse causation in which poor sleep leads to more engagement in R/S practices. For instance, poor sleep may increase emotional distress, leading individuals to seek solace through spiritual practice. Alternatively, high-frequency religious engagement may reflect internal struggles, such as guilt, anxiety, or religious doubt, which may disrupt sleep.^37^ Religious coping can involve praying for comfort and hope during trying times, reading religious writings for inspiration, or seeking support from a faith community.^34^ While these behaviors may provide emotional support, they may also occur in the context of ongoing stress, loss of control, or perceptions of divine punishment, which can sustain physiological arousal, negatively affect psychological well-being, and interfere with restorative sleep.^34, 46–48^ The effectiveness of R/S as a coping mechanism may depend on context (e.g., voluntary vs. obligatory practices), the motivation or intent of engagement (e.g., intrinsic or extrinsic), and the emotional tone of the experience (e.g., comfort vs. struggle).^34^

Contrary to expectations, we did not observe significant longitudinal associations between R/S and any of the sleep dimensions. Several explanations may account for this null finding. First, R/S was measured only at baseline, and thus our analysis did not capture religious history or changes in religious or spiritual engagement and experience (e.g., struggle/trauma) over time. R/S is not a static construct and may fluctuate in response to life events, changing stress levels, or personal growth. Second, it is possible that R/S exerts short-term or immediate effects on sleep that do not persist or accumulate over longer periods. Third, participants may have experienced shifts in their stress exposure or sleep health over time, introducing additional variability that our baseline R/S indicators could not capture. Additionally, the lack of longitudinal associations could reflect the complex and multifactorial nature of sleep, which is influenced by numerous time-varying factors, such as employment and health conditions. Thus, single-time-point measures may not adequately reflect the complex associations between R/S and sleep health, especially if ceiling-effects are present in the measurement of R/S in African-American women.

Our findings should be interpreted in the context of several limitations. First, although we used validated self-reported sleep measures, objective sleep data (e.g., actigraphy) were not available, which may have introduced measurement error or recall bias. Similarly, R/S was assessed only at baseline using limited items that did not distinguish between organizational, non-organizational, and intrinsic religiosity, nor did they capture religious coping styles (e.g., positive vs. negative coping) or experiences of religious struggle. Religious service attendance patterns were also not assessed, so we could not examine attendance-specific associations or compare our findings with the limited existing evidence on sleep health.^49^ These nuances may be important in shaping how R/S influences sleep health. Future studies should include multidimensional and longitudinal assessments of R/S to better understand its dynamic role in sleep health over time and to address the potential for reverse causation, such as the possibility that sleep problems may influence engagement in religious or spiritual practices rather than result from them. The distinctions between religion and spirituality should also be examined as they could have differential impacts on sleep health. Second, our measure of stress was a single-item assessment of perceived day-to-day stress, which, while meaningful, may not fully capture the complexity or chronicity of stress experiences among Black/AA women. Inclusion of validated multidimensional stress scales or biomarkers of stress (e.g., cortisol, allostatic load) in future studies could help clarify the biopsychosocial mechanisms linking stress, R/S, and sleep. We also did not capture detailed childcare and other caregiving responsibilities (e.g., number/ages of children, nighttime caregiving, partner support), which may confound the observed R/S–sleep associations despite adjustment for marital status and employment. Third, the sample included young Black/AA women from Detroit, Michigan, which may limit generalizability to other populations or age groups. However, this demographic is often underrepresented in sleep and health disparities research, and our study helps fill a critical gap by focusing on early adulthood, a life stage when chronic sleep problems often emerge. Despite these limitations, this study has notable strengths. We leveraged data from a large, well-characterized cohort of Black/AA women with repeated assessments over time. The inclusion of multiple sleep dimensions and the consideration of both main along with modifying effects of R/S and stress offers a nuanced understanding of the role of R/S in sleep health.

Our findings underscore the importance of considering culturally salient coping mechanisms, such as R/S, when studying sleep health disparities. While public health and clinical sleep interventions often focus on behavioral and cognitive strategies, integrating the cultural and spiritual values of target populations may improve acceptability and effectiveness, particularly for Black/AA women. For example, faith-based sleep health promotion efforts or spiritually integrated cognitive behavioral therapy for insomnia ^50^ may resonate more deeply within communities for whom R/S plays a central role.^51^ Future research should investigate the role of religious coping styles, religious struggle/trauma, and spiritual support networks in shaping sleep health over time. Longitudinal and qualitative studies may help clarify whether certain forms of R/S serve as proactive coping mechanisms or reactive responses to distress, and how this distinction influences sleep outcomes. Moreover, interdisciplinary approaches that bridge epidemiology, psychology, theology, and community engagement are warranted to develop holistic, equity-informed interventions.

In conclusion, this study highlights the complex and context-dependent relationship between R/S and sleep health among young Black/AA women. R/S as a source of strength was marginally associated with a higher prevalence of restorative sleep among women experiencing high levels of stress, suggesting a potential stress-buffering effect. In contrast, frequent prayer or meditation was associated with NRS, possibly reflecting reactive coping in response to distress. No longitudinal associations were found, underscoring the need for future studies with repeated measures of R/S and more granular assessments of stress and coping. Objectively measured sleep is also warranted. These findings ultimately underscore the importance of incorporating salient, culturally relevant psychosocial factors into sleep health research to better inform interventions aimed at reducing disparities and improving well-being in Black/AA communities.

## Data Availability

The datasets analyzed during the current study are not publicly available due to privacy concerns; however, requests for data may be made by emailing. Data use is restricted to replication, only. New analyses must be approved after a proposal process.

## ACKNOWLEDGEMENTS

The authors would like to thank the participants of the Study of Environmental, Lifestyle and Fibroids.

## AUTHOR CONTRIBUTIONS

*Authors:* Rupsha Singh, Symielle A. Gaston, Jason Ashe, Quaker E. Harmon, Yusuf Ransome, Ganesa Wegienka, Donna D. Baird, Harold G. Koenig, Chandra L. Jackson.

*Study concept and design:* Chandra L. Jackson. *Acquisition of data:* Donna D. Baird, Ganesa Wegienka. *Statistical Analysis:* Rupsha Singh.

*Interpretation of data:* Rupsha Singh, Symielle A. Gaston, Jason Ashe, Quaker E. Harmon, Yusuf Ransome, Ganesa Wegienka, Donna D. Baird, Harold G. Koenig, Chandra L. Jackson.

*Drafting of the manuscript:* Rupsha Singh, Symielle A. Gaston, Jason Ashe.

*Critical revision of the manuscript for important intellectual content:* Rupsha Singh, Symielle A. Gaston, Jason Ashe, Quaker E. Harmon, Yusuf Ransome, Ganesa Wegienka, Donna D. Baird, Harold G. Koenig, Chandra L. Jackson.

*Administrative, technical, and material support:* Chandra L. Jackson.

*Obtaining funding and study supervision:* Donna D. Baird, Chandra L. Jackson.

*Final Approval:* Rupsha Singh, Symielle A. Gaston, Jason Ashe, Quaker E. Harmon, Yusuf Ransome, Ganesa Wegienka, Donna D. Baird, Harold G. Koenig, Chandra L. Jackson.

